# Discovering latent subtypes of preterm birth and genetic risk using tensor decomposition on electronic health records

**DOI:** 10.1101/2025.11.17.25340397

**Authors:** Abin Abraham, Cosmin Bejan, Hannah Takasuka, Marina Sirota, John A. Capra

## Abstract

Preterm birth is a syndrome that is triggered by diverse biological pathways and presents with many comorbid diseases. Although twin studies reveal a substantial heritable component, the genetic mechanisms of preterm birth remain poorly understood. We hypothesize that refining the preterm birth phenotype will reveal sub-phenotypes associated with distinct genetic risk factors and potential treatments. Here, we leverage rich longitudinal data from electronic health records (EHRs) from over 60,000 individuals from two clinical sites. Using tensor decomposition, we uncover several latent factors (LFs) that capture coherent combinations of comorbidities (e.g., metabolic, inflammatory, and mental health) and temporal trajectories of preterm and term births. Similar LFs are discovered between the two sites, underscoring their interpretability. Machine learning models trained on LFs accurately predict preterm birth and perform comparably to models trained on the full EHR data. Integrating genome-wide genotyping for >2,200 individuals, we find robust associations of preterm birth risk with high polygenic burden for cardiovascular disease, type 2 diabetes and body mass index. Using LFs, we discover that these genetic signals are strongly and specifically associated with different subsets of the preterm birth cohort. For example, the polygenic diabetes risk is associated with a LF characterized by relevant metabolic disorders. In summary, our study integrates latent phenotypes discovered from large EHR datasets with genetic data to predict preterm birth risk, uncover disease subtypes and comorbidities that drive genetic associations, and delineate the mechanisms underlying the heterogeneity of this complex trait.

## INTRODUCTION

Preterm birth affects approximately 10% of pregnancies^1,2^ and leads to the highest rates of infant mortality worldwide^3,4^. Researchers have yet to understand the precise mechanisms governing birth timing^5,6^. Many lines of evidence suggest a strong genetic basis for birth timing. For example, family history and previous preterm birth are the strongest risk factors for preterm birth^1^. Additionally, twin-based studies estimate heritability between 17-34%^7,8^. Despite evidence of a strong genetic basis, genome-wide association studies (GWASs) have had limited success in identifying and replicating genomic regions associated with preterm birth^9–11^.

For genetic and mechanistic studies, the phenotypic heterogeneity of preterm birth^5,12^ can limit the power to detect distinct etiologies^13,14^. Preterm birth is often divided into those with spontaneous onset of labor or medically-indicated deliveries. However, this dichotomy does not consider the varied clinical presentation and comorbidities that influence preterm birth risk. A majority of preterm birth cases present with at least one complicating maternal or fetal condition^12^. In addition to obstetric factors, chronic and systemic comorbidities, such as diabetes or mental health disorders, also increase the risk of preterm birth^15^.

A refined definition of preterm birth phenotypes has the potential to identify specific disease mechanisms. For example, the etiology of some preterm birth cases can be narrowed down to abnormal cervical remodeling^16^. Even in a small cohort (n∼100), hierarchical clustering of obstetric variables identified a subset of preterm birth cases associated with the *INS* (insulin) gene^17^.

Electronic health records (EHRs) are a powerful resource for parsing the phenotypic complexity of preterm birth. EHRs are cost-effective and efficient for assembling large cohorts with broadly sampled disease traits across multiple timepoints. Indeed, leveraging disease traits from EHRs has identified novel genotype-phenotype associations^18^ and resolved heterogeneity in sepsis ^19^, diabetes^20^, and cardiovascular disease^21^. EHR-based analyses have recently enabled the prediction of preterm birth risk^22^ and the identification of distinct risk factors ^23^ for spontaneous compared to non-spontaneous preterm birth.

In addition to capturing a broad array of diseases, EHRs document the trajectory of diseases over a patient’s life. Tensor decomposition is an unsupervised method that can model longitudinal disease trajectories while simultaneously decomposing a phenotype into latent factors based on comorbidities. Latent factors capture hidden structure based on the relationships between measured variables within a dataset. Applying tensor decomposition to EHR data for cardiovascular disease, Zhao et al.^24^ demonstrated time-evolving trajectories of distinct subtypes associated with differential long-term survival. EHRs are well-suited to investigate phenotypic heterogeneity in pregnancy. During pregnancy, there is heightened clinical surveillance, measurable endpoints, and rich documentation of the patient’s health. Moreover, a patient’s health history can affect the risk for adverse pregnancy outcomes and adverse outcomes during pregnancy can have long-term health consequences.

In this study, we investigate the phenotypic and genetic heterogeneity of preterm birth. In a large pregnancy cohort, we quantify the phenotypic heterogeneity of preterm birth by identifying comorbid diseases in large EHR cohorts. Next, we apply tensor decomposition on longitudinal EHR data, spanning nine years before and after delivery, and across a large clinical phenome. We uncover 33 latent factors and demonstrate that they are interpretable and capture distinct combinations of comorbid and temporal signatures. Using machine learning, we show that latent factor weights can robustly predict preterm birth with comparable performance to models trained on the full EHR billing code data set. We then quantify the genetic heterogeneity of preterm birth in light of the phenotypes defined by the latent factors using polygenic risk scores (PRSs) for comorbid diseases. We demonstrate the power of latent factor based subphenotyping by uncovering distinct polygenic risk associated with relevant latent factors in preterm and non-preterm cohorts. Our study introduces a new model to dissect preterm birth phenotypic heterogeneity. This approach uncovers genetic risk factors for preterm birth and provides evidence for partitioned genetic risk among subphenotypes of preterm birth.

## RESULTS

### Ascertaining large real-world pregnancy cohorts

We assembled a pregnancy cohort from Vanderbilt University’s EHR database (>3.2 million records) by identifying pregnant individuals with at least one delivery (n=38,402, Table 1). We ascertained the delivery type (preterm vs. not-preterm) using delivery-specific billing codes and estimated gestational age when available (n_preterm_=9,793, n_not-preterm_=28,609, Methods). The age at delivery was similar between the two groups (Table 1). Race, as documented in the EHR, revealed that Caucasian, African American, and Hispanic populations were the most common. We observed a higher proportion of preterm birth in our dataset (25.5%) and within each race category compared to national estimates. This is likely due to sampling from a tertiary healthcare system enriched for preterm births.

**Table 1:** Demographic characteristics of the Vanderbilt pregnancy cohort. From the EHR database, we identified pregnant individuals with deliveries using billing codes and estimated gestational age as available (Methods). The delivery type (preterm vs not-preterm) was ascertained for the earliest delivery in that individual’s EHR. Race for each individual was extracted from the EHR.

|  | not preterm<br>(N=28609) | preterm<br>(N=9793) | Overall<br>(N=38402) |
| --- | --- | --- | --- |
| **Age at Delivery (yrs)** |  |  |  |
| Mean (SD) | 28.8 (5.80) | 28.2 (6.24) | 28.7 (5.92) |
| Median [Min, Max] | 28.9 [13.0, 56.4] | 28.0 [11.9, 55.6] | 28.7 [11.9, 56.4] |
| **Race** |  |  |  |
| CAUCASIAN | 17066 (59.7%) | 6041 (61.7%) | 23107 (60.2%) |
| AFRICAN_AMERICAN | 4813 (16.8%) | 1951 (19.9%) | 6764 (17.6%) |
| HISPANIC | 3499 (12.2%) | 842 (8.6%) | 4341 (11.3%) |
| UNKNOWN | 1315 (4.6%) | 572 (5.8%) | 1887 (4.9%) |
| ASIAN | 1450 (5.1%) | 264 (2.7%) | 1714 (4.5%) |
| OTHER | 362 (1.3%) | 94 (1.0%) | 456 (1.2%) |
| NATIVE_AMERICAN | 75 (0.3%) | 16 (0.2%) | 91 (0.2%) |
| AFRICAN_AMERICAN,CAUCASIAN | 14 (0.0%) | 9 (0.1%) | 23 (0.1%) |
| ASIAN,CAUCASIAN | 8 (0.0%) | 4 (0.0%) | 12 (0.0%) |
| NATIVE_AMERICAN,CAUCASIAN | 3 (0.0%) | 0 (0%) | 3 (0.0%) |
| ASIAN,AFRICAN_AMERICAN | 2 (0.0%) | 0 (0%) | 2 (0.0%) |
| ASIAN,AFRICAN_AMERICAN,NATIVE_AMERICAN | 1 (0.0%) | 0 (0%) | 1 (0.0%) |
| AFRICAN_AMERICAN,NATIVE_AMERICAN | 1 (0.0%) | 0 (0%) | 1 (0.0%) |

To complement the Vanderbilt data, we also analyzed a pregnancy cohort (n=2,295 preterm, not-preterm=20,758, Table 2) from the University of California, San Francisco (UCSF), a tertiary healthcare system with similar EHR data. We identified individuals’ demographics and first delivery type with the UCSF clinician-curated perinatal database and extracted comorbidity data from the EHR through phecodes. The age at delivery was similar between the preterm and term (Table 2). Race, as documented in the perinatal database, revealed that White, Asian/Pacific Islander, and Latina populations were the most common. Compared to the United States, the preterm birth rate in this cohort is similar; however, this population was generally older, more educated, and was privately insured at a high rate.

**Table 2:** Demographic characteristics of the UCSF pregnancy cohort. We identified pregnant individuals’ first deliveries from the clinician-curated UCSF perinatal database (Methods). From this database, we obtained the delivery type (spontaneous preterm, indicated preterm, term), maternal recorded race, maternal education level, and insurance type.

|  |  | spontaneous | indicated | all preterm | term | overall |
| --- | --- | --- | --- | --- | --- | --- |
|  | total (n) | 1129 | 1166 | 2295 | 20758 | 23053 |
| race, n (%) | Single race-White | 467 (41) | 411 (35) | 878 (38) | 9413 (45) | 10291 (45) |
|  | Single race-Black | 75 (7) | 104 (9) | 179 (8) | 1070 (5) | 1249 (5) |
|  | Single race-Latina | 154 (14) | 221 (19) | 375 (16) | 1725 (8) | 2100 (9) |
|  | Single race-Asian/Pacific Islander | 230 (20) | 191 (16) | 421 (18) | 5358 (26) | 5779 (25) |
|  | Single race-Native | 2 (0.2) | 4 (0.3) | 6 (0.3) | 39 (0.2) | 45 (0.2) |
|  | Mul race-Latina + other race | 54 (5) | 60 (5) | 114 (5) | 926 (5) | 1040 (5) |
|  | Mul race-other races | 22 (2) | 22 (2) | 44 (2) | 514 (3) | 558 (2) |
|  | Other race | 54 (5) | 70 (6) | 124 (5) | 1103 (5) | 1227 (5) |
|  | Unknown | 71 (6) | 83 (7) | 154 (7) | 610 (3) | 764 (3) |
| Maternal Age | median | 33 | 33 | 33 | 34 | 34 |
|  | [min,max] | [15,54] | [15,51] | [15,54] | [14,55] | [14,55] |
|  | mean (SD) | 32 (6) | 33 (6) | 33 (6) | 33 (5) | 33 (5) |
| Private Insurance, n (%) | Yes | 915 (81) | 897 (77) | 1812 (79) | 18307 (88) | 20119 (87) |
|  | No | 209 (19) | 263 (23) | 472 (21) | 2377 (12) | 2849 (12) |
|  | unknown | 5 (0.4) | 6 (0.5) | 11 (0.5) | 74 (0.4) | 85 (0.4) |
| Mat. Education, n (%) | 12th_grade | 330 (29) | 434 (37) | 764 (33) | 3563 (17) | 4327 (19) |
|  | college | 634 (56) | 486 (42) | 1120 (49) | 15619 (75) | 16739 (73) |
|  | <12th_grade | 61 (5) | 72 (6) | 133 (6) | 263 (1.3) | 396 (1.7) |
|  | unknown | 104 (9) | 174 (15) | 278 (12) | 1313 (6) | 1591 (7) |
| phecodes per patient | median | 13 | 15 | 14 | 13 | 13 |
|  | [min,max] | [1,91] | [1,135] | [1,135] | [1,124] | [1,135] |
|  | mean (SD) | 16 (11) | 20 (15) | 18 (14) | 15 (10) | 15 (10) |

### Preterm birth is associated with many traits across disease systems

To build a phenome-wide map of traits associated with preterm birth, we mapped each individual’s ICD (International Classification of Diseases) codes to phecodes, which enables us to define their case/control status for thousands of clinically meaningful conditions^25^. With the Vanderbilt cohort, we regressed delivery type (preterm vs. not-preterm) on each phecode with at least four instances per individual while adjusting for age at first EHR-delivery and length of EHR. Women with at least one preterm birth were cases and all others were controls. Given the large disparity in preterm birth rates between African American and Caucasian individuals^26^, we analyzed the two largest EHR-based cohorts of Caucasian (n_cases_ = 6,038, n_controls_ = 16,975) and African American (n_cases_ = 1,951, n_controls_ = 4,805) separately. In these stratified analyses, we considered phecodes to be statistically significantly associated with preterm birth if they pass Bonferroni multiple testing correction (p_Caucasian_< 3.8e-5, p_African American_ < 5.2e-5). The thresholds are different given the different number of testable phecodes in each group.

In the African American cohort, we discovered 28 phecodes significantly associated with preterm birth out of 926 tested (Figure 1A, Table S1). Six out of 17 phecode disease chapters had at least one association with preterm birth. As expected, the pregnancy chapter had the largest number of associations (n=11; Figure 1B, Table S1). Amniotic cavity abnormalities, which include oligo/poly-hydramnios, premature rupture of membranes, infection of amniotic membranes, or spontaneous/artificial rupture of membranes, had the strongest association (p=3.1e-76, OR=5.6). Other preterm birth associated phecodes included hypertensive disorders (preeclampsia p=8.2e-55, OR=5.3; hypertension complicating pregnancy, p=1.9e-26, OR=2.5), cervical incompetence (p=5.2e-26, OR=13.4), and misscarriage (p=2.3e-15, OR=3.6).

**Figure 1.**
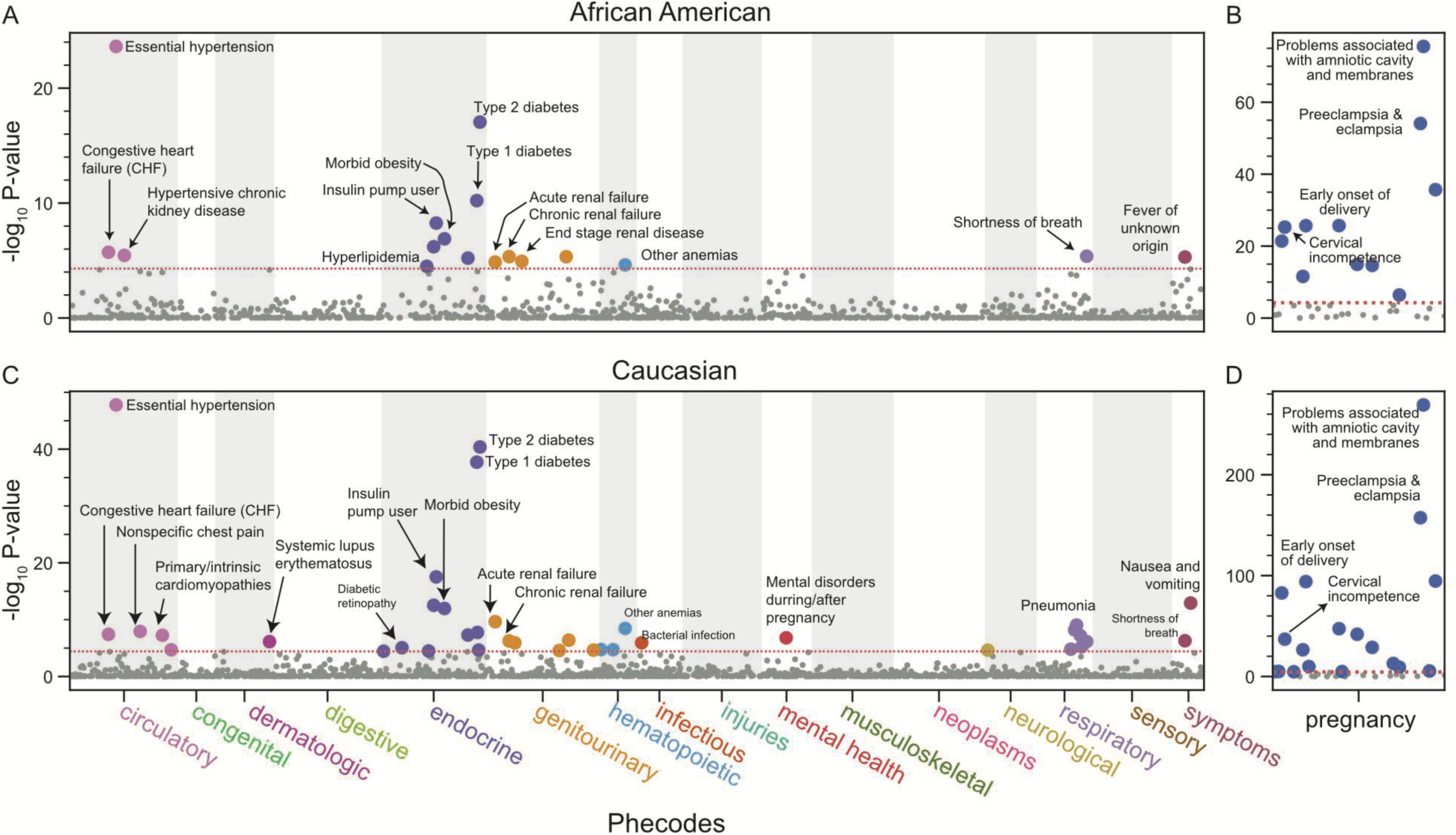
Preterm birth is associated with multiple disease phenotypes across the clinical phenome. We tested for an association between preterm birth and disease phenotypes (captured as phecodes, x-axis) in a cohort of (A, B) African American (n_cases_ = 1,951, n_controls_ = 4,805) and (C, D) Caucasian (n_cases_ = 6,038, n_controls_ = 16,975) women while adjusting for age at delivery and the length of their EHR. Cases included women with at least one preterm birth while controls included women with term or post-term deliveries. Phenotypes are organized by disease category (x-axis) with pregnancy phenotypes plotted separately (B, D) and the negative log_10_ P-value (y-axis). Diseases were considered associated if they passed a Bonferroni correction for number of traits tested (red dotted line) within each cohort (p_African American_ < 5.2e-5, p_Caucasian_< 3.8e-5, Table S1,S2)

In the non-pregnancy disease chapters, the top associations with increased preterm birth risk included known risk factors, including hypertensive traits (essential hypertension, p=2.3e-24, OR=3.4; hypertensive chronic kidney disease, p=3.5e-6, OR=12.5), diabetes (type 2, p=8.9e-18, OR=3.4; type 1, p=6.1e-11, OR=5.9), and morbid obesity (p=1.2e-7, OR=2.0). Many renal traits were also strongly associated with preterm birth (end stage renal disease, p=1.1e-5, OR=11.1; chronic renal failure, p=4.6e-6, OR=12.1; proteinuria, p=6.1e-6, OR=6.4).

In the larger Caucasian cohort, we detected more preterm birth associated phecodes (n=55) compared to the African American cohort, likely due to greater statistical power (Figure 1C,D). The top associations were similar across both cohorts (Table S2). Anemia during pregnancy (p = 9.4e-6, OR=2.9), infection of the genitourinary tract (p=1.3E-5, OR=2.9), and rhesus isoimmunization (p=3.7e-06, OR=2.9) were associated only in the Caucasian cohort. In the non-pregnancy chapters, pneumonia (p=8.8, OR=3.8), mental disorders during/after pregnancy (p = 1.6e-7, OR=1.8), and lupus (p= 7.3e-7, OR=3.2) were only associated with preterm birth in the Caucasian cohort.

### Tensor decomposition enables discovery of longitudinal sub-phenotypes

The strong associations with preterm birth for distinct traits across multiple organ systems supports the syndromic nature of preterm birth and suggests that defining preterm birth as a composite of sub-phenotypes based on similar morbidity could improve our understanding of its phenotypic heterogeneity. The field of tensor decomposition provides methods for decomposing complex multi-dimensional data into combinations of coherent one-dimensional “factors” that capture dominant patterns of variation. To test for the presence of interpretable preterm birth subtypes with distinct phenotypic and longitudinal signatures, we applied tensor decomposition to the trajectories of phecodes around delivery for each individual (Figure 2).

**Figure 2:**
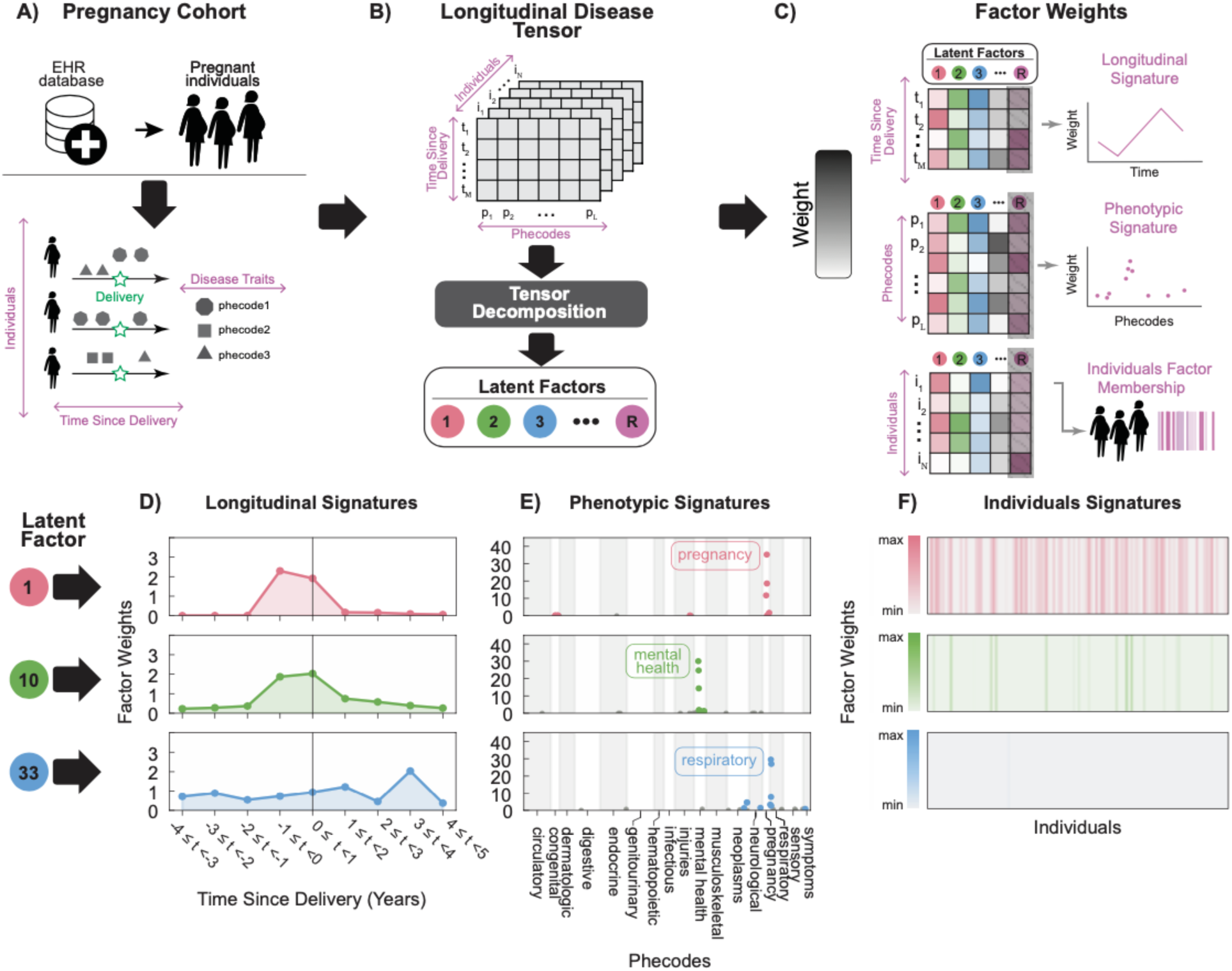
Tensor decomposition identifies latent longitudinal phenotypes within pregnancy cohorts. (A) From the longitudinal EHRs, we generated birth cohorts (n_Vand_=37,068, n_UCSF_=23,356) of pregnant individuals with at least one delivery and disease traits (captured with phecodes) with timestamps. For each individual, we bin the number of occurrences of phecodes into one year temporal bins before and after delivery for the first pregnancy. (B) A 3-D tensor is created by concatenating along the third dimension each individual’s 2-D matrix of phecodes by time since delivery. After performing tensor decomposition, we identify a number of latent factors that capture the underlying structure in the input tensor. (C) Tensor decomposition yields loading matrices with weights for each axis of the 3-D tensor across all latent factors. Plotting weights for a latent factor across time since delivery, phecodes, and individuals describes longitudinal, phenotypic, and individual membership signatures respectively. (D,E,F) We illustrate this by selecting three latent factors (1, 10, 33) from the Vanderbilt cohort with distinct signatures based on the factor weights (y-axis). (D) Time since delivery (x-axis) with negative and positive numbers indicating before and after delivery respectively. (E) phecodes arranged by disease categories (x-axis). (F) Each vertical strip represents one individual with color intensity representing the factor weight.

For each individual in each pregnancy cohort (Vanderbilt and UCSF), we calculated the time since the index delivery (defined as the first EHR-delivery) of phecodes occurring up to four years before and five years after the first EHR delivery (Figure 2A). We structured this dataset into a 3-D tensor in which one dimension represents individuals, one dimension represents phecode occurrence, and the final dimension represents time. We summed each individual’s phecodes in temporal one-year bins from the index delivery. We then applied CANDECOMP/PARAFAC tensor decomposition on this 3-D tensor (Figure 2B). To select the number of latent factors, we maximized between factor uniqueness and within factor coherence (Methods, Supplemental Figure 1). Each of the 33 (at Vanderbilt) or 26 (at UCSF) resulting latent factors is defined by its weights for each mode of the 3-D tensor. This analysis yielded three latent factor weight matrices: time-since delivery, phecodes, and individuals (Figure 2C). Each individual can be represented by their weights for the latent factors in each of these dimensions.

To illustrate the latent factors, we highlight three (latent factor 1, 10, 33) with distinct longitudinal phenotypic signatures (Figure 2D,E,F). Latent factors 1 and 10 capture peri-delivery temporal signatures with their peaks occurring within one year before and after delivery respectively. In contrast, latent factor 33 demonstrates chronic temporality with a peak between three and four years after delivery (Figure 2D). The three latent factors 1,10, and 33 also have distinct phenotypic signatures dominated by pregnancy, mental health, and respiratory traits respectively (Figure 2E). Finally, the weight matrix across individuals and latent factors demonstrates that many individuals have high weights for latent factor 1, and a different, smaller subset of individuals have higher weights for latent factor 10, followed by a small number of individuals with high weights for latent factor 33 (Figure 2F).

To summarize the patterns represented by the latent factors, we examined the highest weight components per phecode category (Figure 3). As expected, for the first 10 latent factors, several have high weights for combinations of pregnancy phecodes at Vanderbilt (1, 2, 3, 4, 6, Figure 3A) and UCSF (1,2,3,8, Figure 3B). Reflecting their coherence and interpretability, the majority of LFs have high weights for a small number of phenotypes in a single phenotypic chapter. In the temporal domain, almost all factors have high weights for phenotypes between delivery and one year prior. Nonetheless, each latent factor has distinct temporal signatures that vary by maximum weight and by its distribution over the time bins (e.g., acute vs chronic patterns, Supplemental Figure 2A). Across individuals, we found that latent factors 1 and 2 have the largest number of individuals with high weights and some latent factors (LF 17, 19, 24) have few individuals with high weight values (Supplemental Figure 2B). To evaluate the similarity of latent factors across phenotypes, longitudinal signatures and individuals, we calculated the Pearson correlation coefficient on the 3-D tensor for each latent factor. We find low pairwise correlation (Pearson R^2: −0.01 to 0) between latent factors which further validates that each latent factor captures distinct signatures (Supplemental Figure 3).

**Figure 3:**
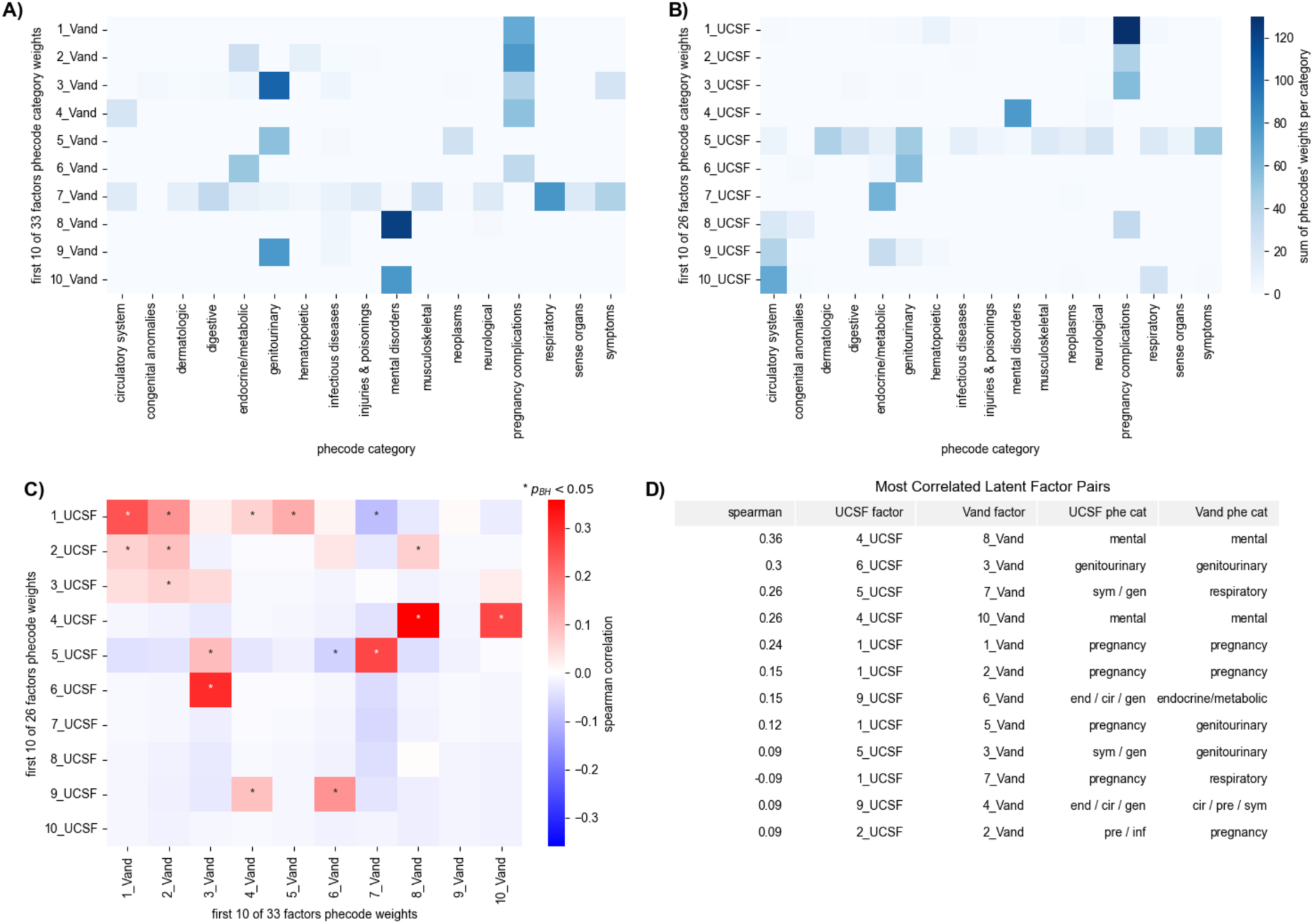
Latent factors identify distinct pregnancy comorbidities and are similar across Vanderbilt and UCSF cohorts. Latent factor weights per phecode category, summarized as the sum of phecode weights for each category at Vanderbilt (A) and UCSF (B). The first 10 factors are shown for brevity. (C) Spearman correlations of phecode weights for each Vanderbilt latent factor with each UCSF latent factor. Asterisks indicate latent factor pairs with significant correlations between their phecode weights after multiple testing correction. (D) Latent factor pairs between Vanderbilt and UCSF with the strongest Spearman correlations. “Phe cat” refers to the major phecode category(s) for the factors listed in each row. One major phecode category is listed if 5 or more of the top 10 phecode weights in that factor fall in the same phecode category. Multiple categories are listed if no category represents at least 5 of the top 10 phecode weights.

### Similar sub-phenotypes are present across pregnancy cohorts

To quantify the similarity of the decomposition across different pregnancy cohorts, we compared latent factor weights at Vanderbilt and UCSF (Figure 3). All but one of the top 10 latent factors at Vanderbilt have a significant correlation with a UCSF latent factor. For example, pregnancy complication phenotypes dominate the first two factors and show significant correlations between institutions (Spearman: 0.06-0.24, p-value benjamini-hochberg corrected <0.05, Figure 3C,D). Vanderbilt’s mental phenotype factor (factor 8) significantly correlates (Spearman=0.36) with UCSF’s mental phenotype factor (LF 4).

Vanderbilt’s genitourinary phenotype factor (factor 3) significantly correlates (Spearman=0.30) at UCSF (LF 6). Vanderbilt factor 7 significantly correlates (Spearman=0.26) with UCSF factor 3, representing phecodes from many categories, including symptoms, genitourinary, and respiratory. Similar to Vanderbilt, most UCSF latent factors have the highest temporal weights between delivery and 1 year prior, and the first few UCSF factors have the largest number of individuals with high weights Supplemental Figure 2C,D).In summary, our unsupervised approach captures similar latent structure at two institutions and provides interpretable longitudinal phenotypic latent factor for pregnancy comorbidities.

### Latent factors perform comparably to phecodes for predicting preterm birth

The weights across latent factors for each individual represent longitudinal comorbid signatures. For most Vanderbilt latent factors their weight distribution across individuals is dominated by very low values with a few outliers (Figure 4A). However, the individual weights for 22 out of the 33 Vanderbilt latent factors (67%) were statistically significantly associated with preterm birth (Figure 4B). Five out of the 22 preterm birth associated Vanderbilt latent factors had negative effect sizes, which suggest a protective latent factor phenotype (Figure 4B). At UCSF, 8 of the 26 latent factors (31%) significantly associated with preterm birth, 3 of which were protective.

**Figure 4:**
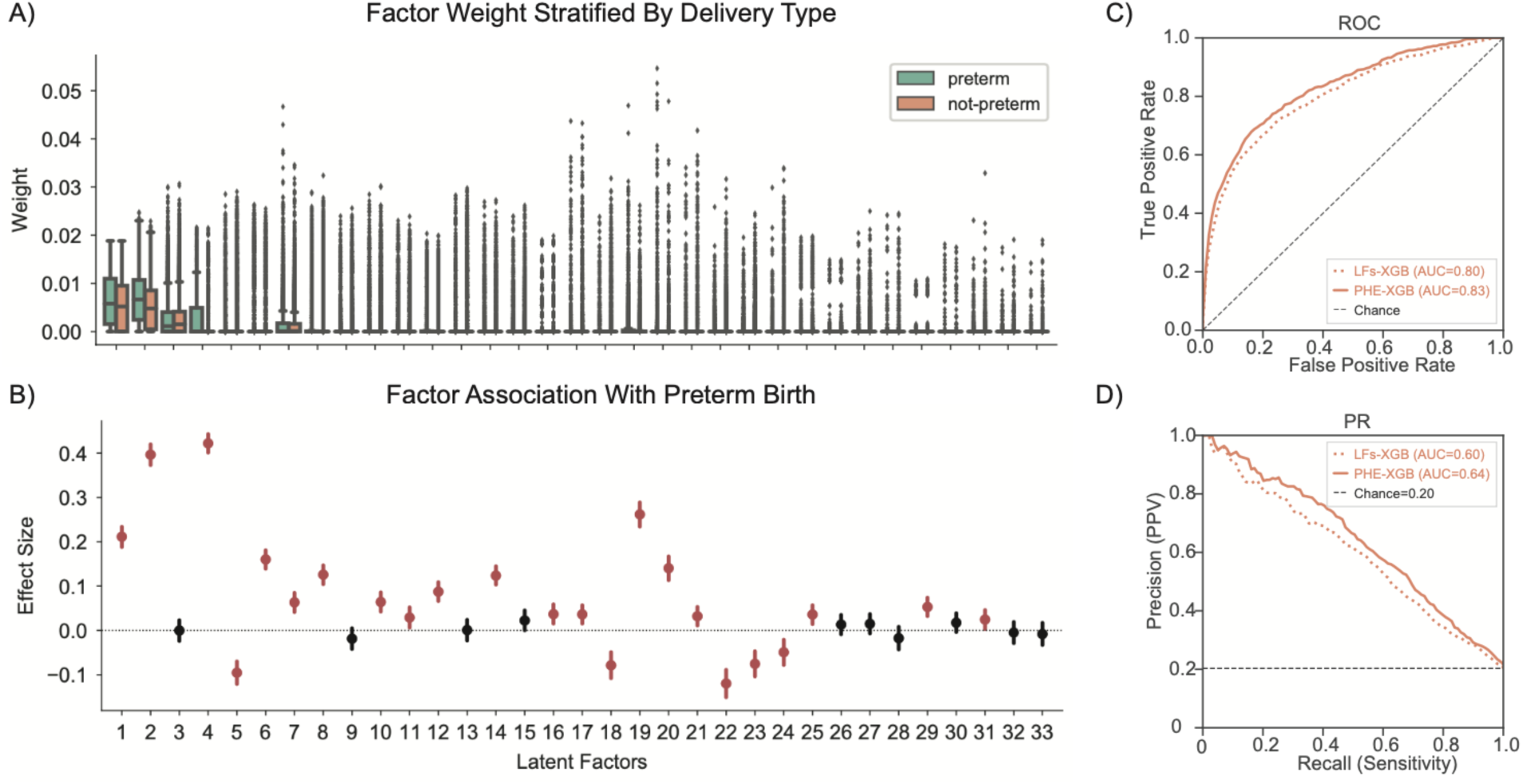
Prediction of preterm birth from latent factors. For each Vanderbilt individual, the phecode weights across the 33 latent factors were used to predict delivery type (preterm vs. not-preterm). (A) Distribution of weights (y-axis) for each individual across latent factors (x-axis) by delivery type. (B) Individual’s weights for many latent factors were significantly associated with delivery type in a logistic regression. Effect size (y-axis) with 2.5 and 97.5th percentile confidence intervals for the association are plotted for each latent factor (x-axis). Associations passing a FDR controlled multiple testing correction (FDR<0.05) are colored red. (C) ROC and (D) PR curves for predicting delivery type using xgboost with latent factor weights (dotted line) and all phecodes (solid line).

Given these associations between individual factors and preterm birth, we tested if they could predict preterm birth at Vanderbilt in a supervised machine learning framework. For comparison, we trained similar models using all phecodes used to create the decomposition as predictors of preterm birth. We have previously shown that phecode models can predict preterm birth with high accuracy^22^. Using boosted decision trees (XGBoost), we found that phecodes (ROC-AUC=0.83, PR-AUC=0.64) and latent factor weights (ROC-AUC=0.80, PR-AUC=0.60) performed similarly in predicting preterm birth (Figure 4C, D). Thus, the latent factors derived from the tensor decomposition maintain most of the information necessary to predict preterm birth.

### Preterm birth is associated with polygenic risk for comorbid traits

Next, we examined whether genetic risk for potentially comorbid traits is associated with preterm birth risk. We downloaded validated polygenic risk scores for various (n=62) potentially comorbid disease traits. For genetic risk analyses, we subset the Vanderbilt pregnancy cohort to include only Caucasian women with genetic data (n=2,288) since the majority of the polygenic risk scores were developed in European ancestry populations. After quality control of the genetic data, we calculated a polygenic risk score for each individual and trait (Methods). Some traits had many validated polygenic risk scores.

Finally, we tested for an association between delivery type (preterm vs not-preterm) and the PRS for each trait while correcting for genetic ancestry (Methods). After multiple testing correction, we discovered nine total associations with PRS scores: body mass index (2 out 5 PRS tested), type 2 diabetes (1 out 7 PRS tested), and coronary artery disease (6 out of 14 tested, Figure 5). All statistically significant associations showed increasing risk for preterm birth with increasing polygenic risk for that trait.

**Figure 5:**
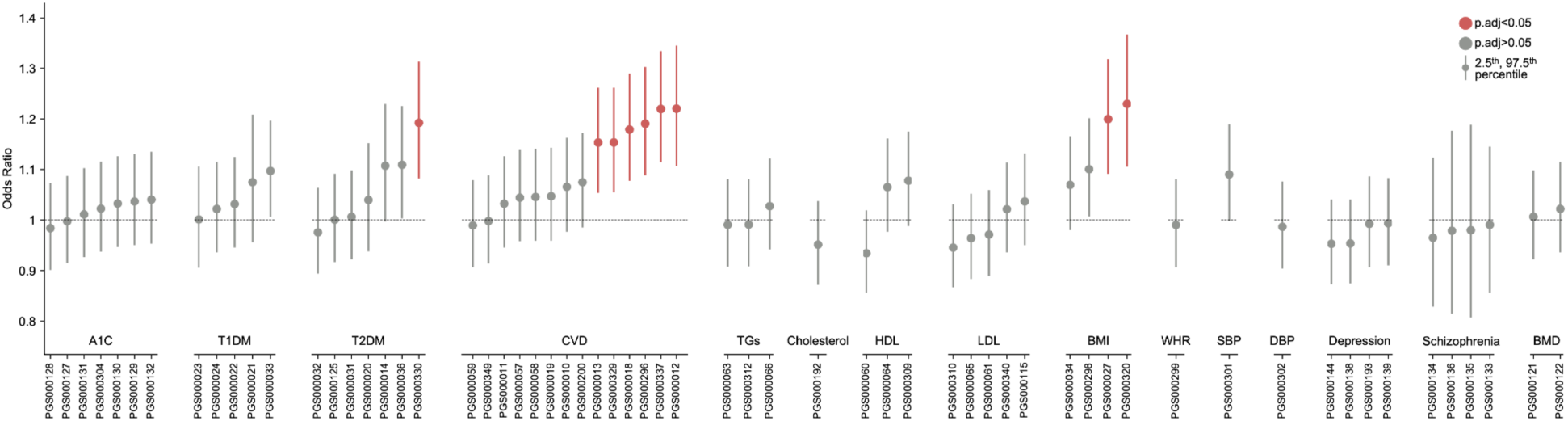
Association of polygenic risk scores with preterm birth. We tested for an association between delivery type (preterm vs. not-preterm) and polygenic risk scores for many traits associated with preterm birth (x-axis, arranged by traits) while controlling for genetic ancestry in the Vanderbilt pregnancy cohort. These analyses focused on Caucasian individuals given the populations in which most PRS were developed. We plot effect size (y-axis) with 2.5th and 97.5th percentile confidence intervals. Associations are colored red if they pass multiple testing correction using false discovery rate across all tests performed.

### Latent factors define subpopulations with different genetic risk factors

The latent factors derived from tensor decomposition partitioned the preterm birth phenotype based on morbidity patterns. Therefore, we hypothesized that the weights of specific latent factors across individuals would be associated with the genetic predisposition for the corresponding comorbidities. To test this with a focus on preterm birth-relevant phenotypes, we split the Vanderbilt cohort into preterm and not-preterm deliveries. Then for each group, we tested the association of each latent factor with polygenic risk scores for comorbid traits, including genetic ancestry as a covariate. We repeated this analysis for each combination of 33 latent factors and 60 polygenic risk scores, which covered 13 major traits (Methods).

Within the preterm birth cohort, we found 6 statistically significant (FDR < 0.05) associations between PRSs and latent factors (Figure 6A). These all indicated that increasing polygenic risk for type 1 and type 2 diabetes were associated with increasing weights for latent factors 6 and 13. Both of these factors were characterized by high weights for metabolic and endocrine phenotypes, such as diabetes (phecode 250), insulin pump user (phecode 250.3), hypoglycemia (phecode 251.3), hypothyroidism (phecode 244), and thyroiditis (phecode 245). This demonstrates that variation in the weights for these latent factors, one of which (factor 6) is associated with preterm birth risk in the entire cohort and one that is not (factor 13), is significantly associated with genetic risk for diabetes. We also found 115 nominally significant (p<0.05) associations between PRSs and latent factors.The majority of these associations were concordant in direction of effect across PRS for a given trait, and most showed similar phenotypic specificity and agreement. For example, PRS for depression and schizophrenia are associated with latent factors dominated by mental health phenotypes. Similarly, within the non-preterm birth cohort, we found 135 nominally significant associations, 7 of which survived multiple testing correction (Figure 6B). These included associations between latent factor 6 and type 1 diabetes and type 2 diabetes, and latent factor 4 with BMI. Comparing the associations between preterm and term cohorts reveals broadly similar patterns. These results show that polygenic risk for comorbid traits, like diabetes, contributes to some, but not all of the phenotypic heterogeneity captured in the latent factors.

**Figure 6:**
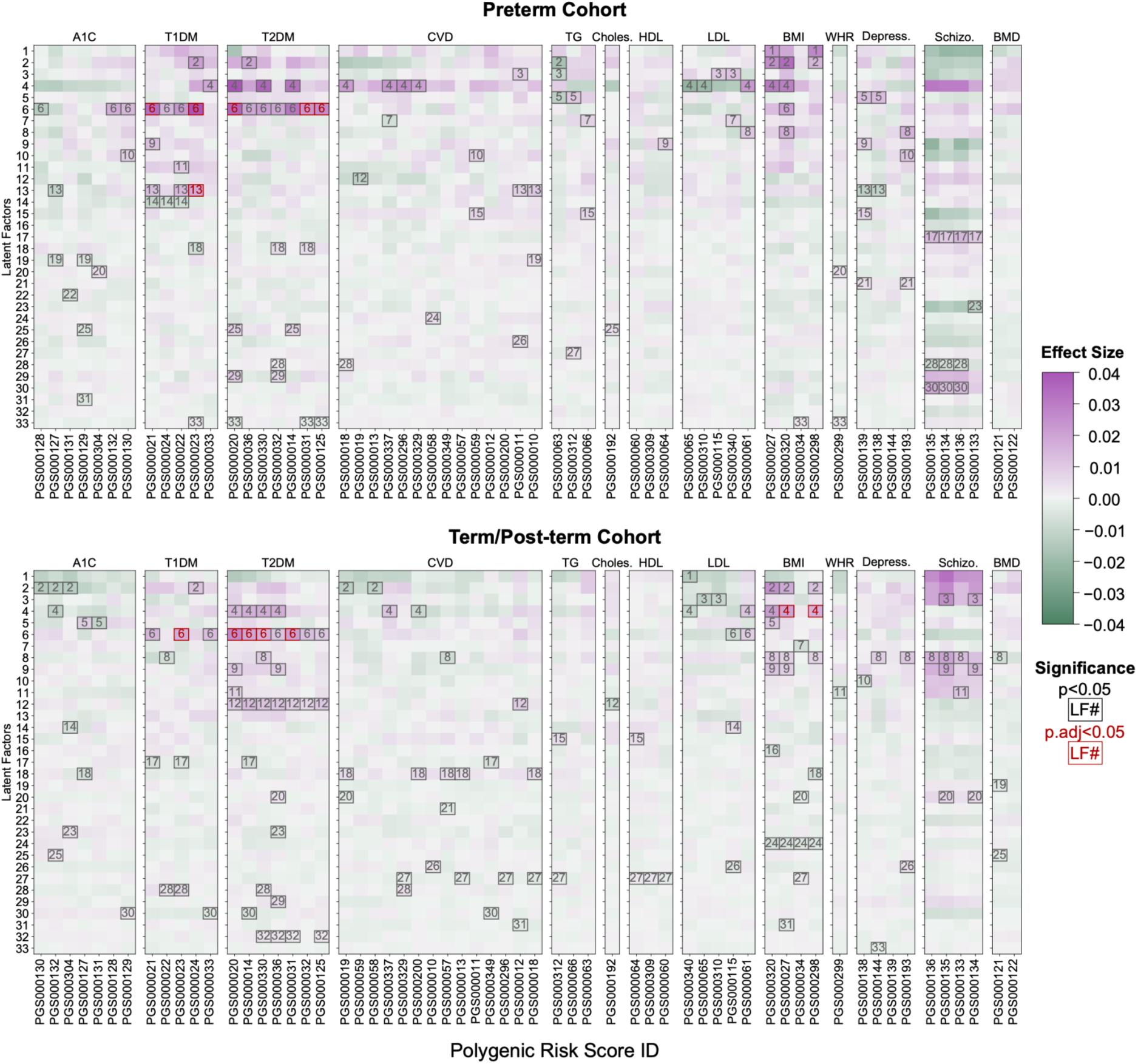
Association of latent factors with polygenic risk scores. For (A) preterm births and (B) term/post-term births at Vanderbilt, we tested for an association between each individual’s weight for a latent factor (rows) and their polygenic risk score for various disease traits (columns). Cells are colored based on their effect-size. For nominal (p<0.05) and multiple testing corrected (FDR<0.05) statistical significant associations are marked in gray or red boxes, respectively.

## DISCUSSION

Despite significant morbidity and mortality, there exists no effective treatment for preterm birth and its underlying mechanisms remain poorly understood. In this study, we tested the hypothesis that analyzing complex real-world clinical data on the preterm birth phenotype could reveal subtypes with distinct genetic risk contributing to birth timing. Leveraging large EHR databases linked to a genetic biobank, we applied tensor decomposition on thousands of disease traits across several years on diverse Vanderbilt and UCSF cohorts to generate latent factors. Each latent factor represents a sub-phenotype distinguished by a unique combination of comorbidities and temporal patterns. Most of the strongest latent factors’ phenotype signatures at Vanderbilt correlate with factors determined on an independent cohort at UCSF. Compared to direct associations of polygenic risk scores and preterm birth, our approach identified additional genetic associations with sub-phenotypes. For instance, PRS for type 1 diabetes was linked to latent factors in preterm and non-preterm cohorts but not with the broader preterm birth phenotype. These findings lend support that specific sub-groups have different genetic and non-genetic risk factors. We hope that precision phenotyping will accelerate our understanding of preterm birth biology.

Applying tensor decomposition to a heterogeneous syndrome like preterm birth revealed its potential to identify known and novel disease subtypes. While similar approaches have been applied to other disease traits^24,27,28^, large scale, phenome-wide, assessment of preterm birth has not yet been performed. Tensor decomposition is flexible in generating latent factors; by applying an orthogonality constraint for phecodes, we obtained latent factors with distinct phenotypic signatures. The decomposition includes longitudinal trajectories linked with phenotypic signatures for each latent factor. By analyzing the distribution of weights across each latent factor for phecodes and time since delivery, we were able to quantify and describe these patterns in finer detail. Compared to other clustering approaches, such as k-means or hierarchical clustering, tensor decomposition links several clinically relevant dimensions together (disease traits, temporality, and individuals). Regression based approach can also quantify preterm birth risk; however, the weights obtained from tensor decomposition enable interpretation and downstream analyses. For example, we exemplify one use case by predicting preterm birth from these weights. Future studies can correlate latent factor weights with other clinical variables of interest.

In addition to refining the phenotypic heterogeneity, we characterize genetic risk factors that contribute to preterm birth. We find that genetic risk for many complex traits (type 2 diabetes, cardiovascular disease, and BMI) also significantly associates with preterm birth risk. While these traits have been linked with increased preterm birth risk in epidemiological studies, our analysis suggests comorbid genetic pathways contributing to birth timing. Of note, our analysis was restricted to Caucasian women since these scores were initially developed in this population and numerous studies have demonstrated the lack of transferability of PRS across human populations^29^. We tested many PRS for the same trait since assigning a ‘best’ PRS is challenging. For each PRS, the initial study population and study design will affect the generalizability of these scores. The PRS for cardiovascular risk was statistically significant after multiple testing correction in six out of 14 PRS that we tested. We hope that this PRS analysis will serve as a proof of concept for investigating how genetic risk from comorbid traits can affect preterm birth risk.

Highlighting a unique strength of latent factors to subgroup preterm birth, we show additional comorbid polygenic risk associated with certain latent factors. For example, the polygenic risk score for type 1 diabetes is significantly associated with certain latent factors in preterm and non-preterm cohorts but not when tested against the complete preterm birth phenotype. This finding points towards subphenotypes with distinct genetic risk contributing to birth timing. The tensor decomposition leverages the inherent structure within the data. Since underlying biological processes manifest as phenotypic correlations that are captured in EHRs, we posit that using tensor decomposition provides an unbiased, first-pass approach to uncover birth timing mechanisms.

Our study relies on EHR data, which has several strengths and limitations that are important to consider. First, EHRs may contain errors in how ICD billing codes are assigned for specific phenotypes. To mitigate this effect, we took several steps: we required multiple instances of a billing code to be present; we mapped billing codes to better defined phecodes;. and we excluded extremely rare codes in our cohort.

Second, missing diagnosis data and misreported race/ethnicity data could produce biases in our dataset. Finally, given that we only study pregnancies at tertiary care centers, our results may not be representative of all preterm births.

Our results motivate several future lines of work. First, the generalizability of our results to other care centers, settings, and populations should be evaluated. Second, we discovered genetic associations with specific latent factors defined in the pregnancy cohorts. Future studies should examine how latent factors and their genetic associations compare between preterm and non-preterm birth cases to improve the ability to predict preterm birth risk. Finally, a detailed dissection of the phenotypes and temporal trajectories in the latent factors associated with preterm birth risk is needed. This has the potential to reveal new causal mechanisms for subtypes of preterm birth.

## METHODS

### Ascertaining pregnancy cohort and delivery type from electronic health records

From the Vanderbilt University Medical Center EHR database of over 3.2 million records, we identified a pregnancy cohort of individuals with at least one delivery at Vanderbilt (n=38,402, Table 1). Using billing codes (ICD-9 and CPT), their timestamps, and estimated gestational ages (EGAs), we applied a validated phenotyping algorithm (precision=0.96, recall=0.99 for preterm birth; 0.99, 0.99 for not-preterm birth) to determine the date and type of each delivery^22^. Delivery codes occurring within 37 weeks of the most recent delivery code were grouped into a single pregnancy, and this process was repeated until all codes were assigned. EGA values within each individual’s EHR were grouped into one pregnancy if they fell within the gestational window indicated by the most recent EGA. Each delivery was classified as preterm or not-preterm (term or post-term deliveries) based on the oldest gestational age captured in the pool of delivery codes and EGAs. We refer to this cohort as the Vanderbilt pregnancy cohort.

At UCSF, we identified a pregnancy cohort using the perinatal database, which is maintained and curated by obstetricians at UCSF, with 46,902 individuals prior to any filtering. The database includes hundreds of demographic and clinical variables about each delivery. We extracted data on maternal age, maternal education level, insurance type, gestational age, and whether the birth is indicated preterm (“medically indicated” or “Termination Iatrogenic”), spontaneous preterm (“spontaneous,” “PPROM,” or “PTL with TOCO and TERM”), or term. We refer to this cohort at the UCSF pregnancy cohort.

### Testing disease traits for association with preterm birth

Using self-reported or third-party documented race in the EHR, we split the Vanderbilt pregnancy cohort into Caucasian and African American cohorts. We labeled individuals with at least one preterm birth as ‘preterm’, while all others were labeled ‘not-preterm’. Next, at Vanderbilt, we queried disease traits across the clinical phenome using billing codes. We extracted all ICD-9/10 codes from each individual’s EHR and translated them to Phecode (version 1.2)^30^. Phecodes group ICD-9/10 codes into clinically relevant categories organized around related diseases. The positive predictive value of ascertaining a disease trait increases when multiple instances of that code appear in a patient’s EHR^25,30^. Thus, we binarized phecodes by requiring at least four instances of a code for an individual to be considered positive for that disease trait. After performing these filtering steps, our dataset included 23,013 Caucasian individuals (preterm=6,038; not-preterm=16,975) and 6,756 African American individuals (preterm=1,951, not-preterm=4,805). We regressed delivery type on each phecode while adjusting for age at first EHR-delivery and length of EHR - defined as the time between the earliest and most recent billing code. This analysis was performed separately in the African American and Caucasian cohorts. Only phecodes occurring in at least 100 individuals were tested for an association with delivery status. To correct for multiple testing, we used the Bonferroni correction over all phecodes tested in each cohort (n_phecodes_=1,306 for Caucasians and 959 for African Americans). ICD to phecode translation and association testing was performed using the PheWAS R package (version 0.99)^31^ using R (version 4.4.0).

### Creating the longitudinal disease tensor from the pregnancy cohort

From the pregnancy cohorts, we selected the earliest pregnancy for each individual. We removed delivery-related billing codes to avoid confounding downstream analyses (Supplement Table S3). For each individual, we categorized the frequency of phecodes into temporal bins. Each bin spanned one-year intervals, covering up to four years before and five years after the first EHR-documented pregnancy. To reduce dataset size, we excluded phecodes present in fewer than 0.5% of individuals. We then created a 3-D tensor by aggregating each individual’s 2-D tensors of phecodes by time since delivery.

The final Vanderbilt tensor dimensions included 1,728 phecodes, 9 temporal bins, and 37,068 pregnant individuals. The UCSF dimensions included 1,277 phecodes, 9 temporal bins, and 23,356 pregnant individuals.

### Tensor decomposition using parallel factor analysis

On the longitudinal disease tensors at each institution, we performed tensor decomposition using parallel factor analysis with alternating least squares. We constrained the factorization to be non-negative across all dimensions and orthogonal in the phecode dimension. After setting the number of latent factors, we used alternating least squares for decomposition until the model fit converged (change in R^2 ≤ 1e-10). For each fixed number of latent factors, we repeated the decomposition twenty times with random initializations and retained the model with the highest R^2. To identify the optimal number of latent factors, we evaluated tensor decompositions with latent factors ranging from three to fifty.

We assessed between phecode-factor independence as the mean Jaccard index of all pairwise factors using weights across the top fifty phecodes. We measured within-factor similarity using the UMass metric, which evaluates the co-occurrence of similar topics, or phecodes, across a set of EHRs. For each factor, we calculated the UMass metric as the sum across the top fifty phecodes with the highest weights. To summarize within-factor similarity, we averaged the summed UMass metric across all factors. To compare Jaccard and UMass metrics on similar scales, we multiplied the UMass metric by −1 and scaled its range between 0 and 1, where higher scores indicated better topic coherence.

Although the UMass metric increased with more latent factors, causing the sum of UMass and Jaccard metrics to rise, the Jaccard index decreased as the number of latent factors increased. To balance UMass and Jaccard metrics while keeping the number of latent factors low for interpretability, we selected 33 latent factors at Vanderbilt and 26 latent factors at UCSF for downstream analyses.

### Predicting preterm birth from latent factors

From the Vanderbilt pregnancy cohort, we used individuals’ weights across latent factors as input features to predict a binary outcome of preterm or not-preterm birth. Delivery-specific phecodes were removed before constructing the longitudinal disease tensor for tensor decomposition. Using a stratified shuffle, we randomly split the data 90:10 with an equal proportion of cases into training and test sets. On the training set, we trained gradient-boosted decision trees and optimized hyperparameters using tree of Parzen estimators (hyperopt v0.1.1)^32^ by maximizing the mean average precision. We evaluated model performance on the test set using standard metrics (PR-AUC, ROC-AUC). The baseline chance performance for precision-recall curves was the prevalence of preterm birth cases. We compared performance of a model based on the latent factors vs. a model trained using the same procedure, but with phecodes as input.

To compare with our gradient-boosted model, we fitted a logistic regression using default ‘L2’ penalty on the training data, assigning sample weights inversely proportional to class frequencies. We evaluated the logistic regression model on the test set and reported standard performance measures.

### Genotyping and quality control

A subset of the Vanderbilt pregnancy cohort including Caucasian pregnant individuals with delivery labels (n=2,288) had genotype data available from the Illumina MEGA-Ex chip (>2 million common and rare SNPs). We followed standard population genetics quality controls, and we excluded variants with a minor allele frequency < 0.01 or a missingness rate > 0.05 using PLINK v1.90b4s^33^. We performed imputation through the Michigan Imputation Server for the European population and removed imputed variants with an INFO score below 0.3^34^.

### Calculating polygenic risk scores and testing for preterm birth association

We applied polygenic risk scores (PRS) for known comorbidities associated with preterm birth in Caucasian pregnant individuals within our cohort. We restricted the analysis to Caucasian individuals because most PRS were developed in European populations. The comorbid traits tested included BMI, hemoglobin A1C, Type 1 Diabetes, Type 2 Diabetes, triglycerides, LDL, HDL, cholesterol, cardiovascular disease (CVD), coronary artery disease, systolic and diastolic blood pressure, depression, schizophrenia, and bone mineral density. For each trait, we downloaded SNP weights for validated PRS from the Polygenic Risk Score Catalog (PGS, version 1)^35^.

We calculated the PRS for each individual as the sum of risk alleles multiplied by the weights from the PGS. This analysis was performed using the --score function in PLINK v1.90b4s. After normalizing the PRS for each trait, we tested for associations using logistic regression, with delivery type (preterm vs. not-preterm) as the dependent variable and the PRS for each trait as the independent variable. A pregnant person with at least one preterm birth was classified as ‘preterm’ (n_preterm_=890), while all others were labeled ‘not-preterm’ (n_not-preterm_=1398). Covariates included genetic ancestry with 15 principal components. We adjusted p-values for multiple testing using the Benjamini/Hochberg method and considered an adjusted p-value < 0.05 as significant. All analyses were performed using the statsmodels (version 0.11.1) Python package.

### Testing for association between latent factor and polygenic risk scores

We applied multiple linear regressions to test for associations between latent factor weights and PRS. This analysis was conducted separately for preterm and not-preterm birth cohorts derived from our pregnancy cohort. Latent factor weights served as the dependent variable, while polygenic risk scores were the independent variable. We included 15 genetic ancestry principal components as covariates.

For each regression model, latent factor weights were normalized, and PRS were z-score standardized by subtracting the mean and dividing by the standard deviation. This approach enabled comparison across latent factors and PRS. For all associations tested, increasing PRS indicated greater risk for the trait. We report the effect size and its confidence intervals (2.5th, 95th percentiles). For both preterm and not-preterm cohorts, we corrected for multiple testing by adjusting p-values using the Benjamini/Hochberg method (statsmodels, v0.11.1).

## Data Availability

All computational methods and code are provided via a Github repository (https://github.com/abraham-abin13/ptb_latent_factor). Individual level data is not provided due to privacy concerns associated with electronic health records, but can be obtained via application from the Vanderbilt University’s BioVU program and UCSF Information Commons. Public datasets used include polygenic risk score weights obtained from the The Polygenic Score (PGS) Catalog.

## ACKNOWLEDGEMENTS

We thank the members of the Capra lab for thoughtful discussions on this project.

## CONTRIBUTIONS

*Conceptualization and methodology*: A.A. and J.A.C. conceived and designed the study. *Data curation & Resources*: A.A. and C.A.B. extracted the billing codes for the Vanderbilt cohort and curated publicly available polygenic risk scores. H.T. extracted the billing codes for the UCSF cohort. *Formal analysis and investigation*: A.A. performed all analyses on the Vanderbilt cohort; H.T. carried out all analyses on the UCSF cohort. J.A.C. supervised all analyses. Funding acquisition: J.A.C. Writing: A.A. wrote the manuscript with guidance from H.T., M.S., and J.A.C.

## SUPPLEMENTAL FIGURES

**Supplemental Figure 1:**
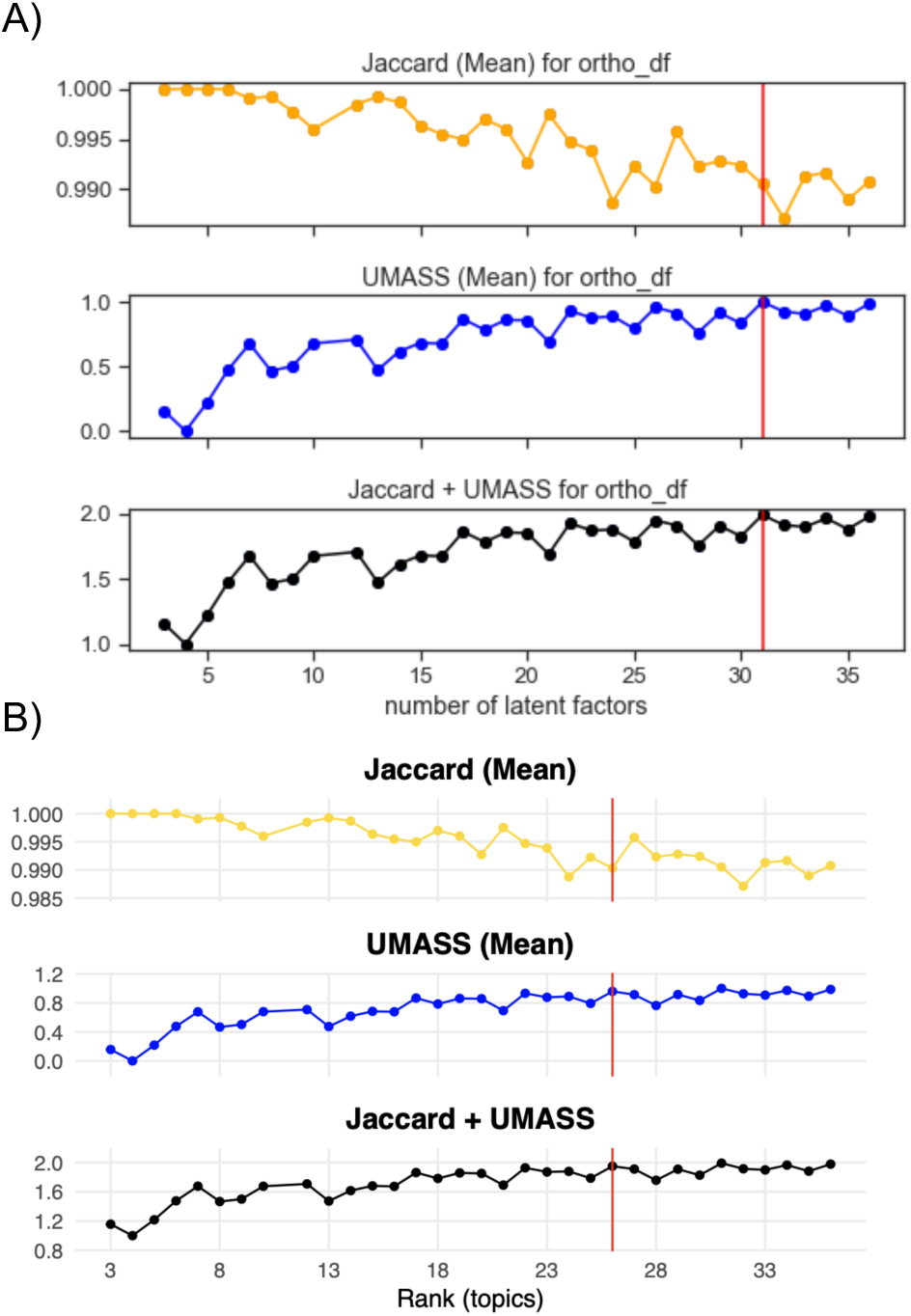
Optimal Latent Factor Metrics (A) Vanderbilt (B) UCSF. Across different number of latent factors (x-axis), the top panel plots the mean Jaccard index (y-axis) of all pairwise factors using weights across the top fifty phecodes, middle panel plots the within-factor similarity using the UMass metric, and the bottom panel is the sum of the top two panels. The red line indicates the optimal number of latent factors (33 for Vanderbilt and 26 for UCSF). See Methods for details.

**Supplemental Figure 2:**
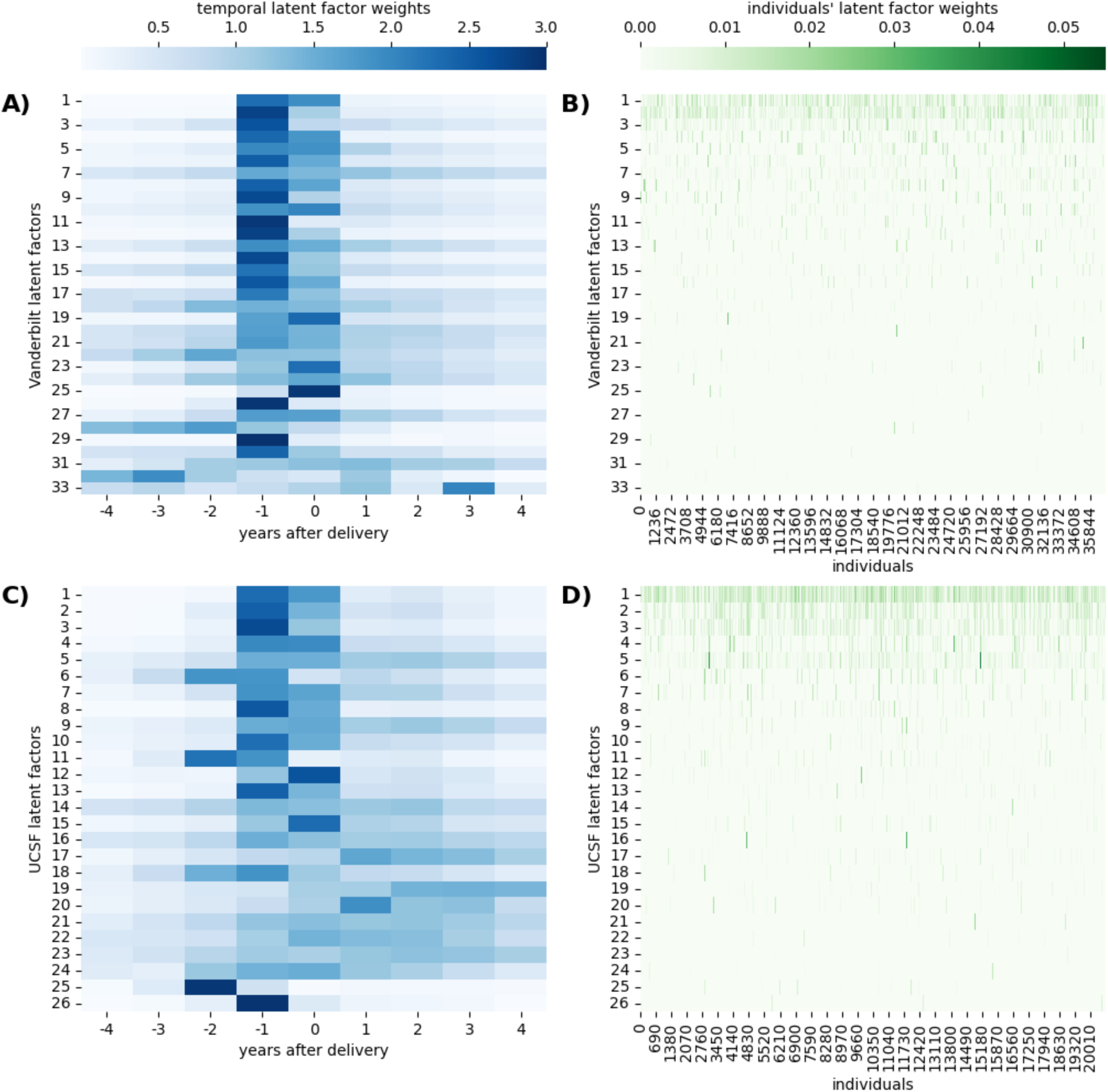
Latent Factor Weights per temporal bin (A, C) and individual (B, D) at Vanderbilt (top) and UCSF (bottom).

**Supplemental Figure 3:**
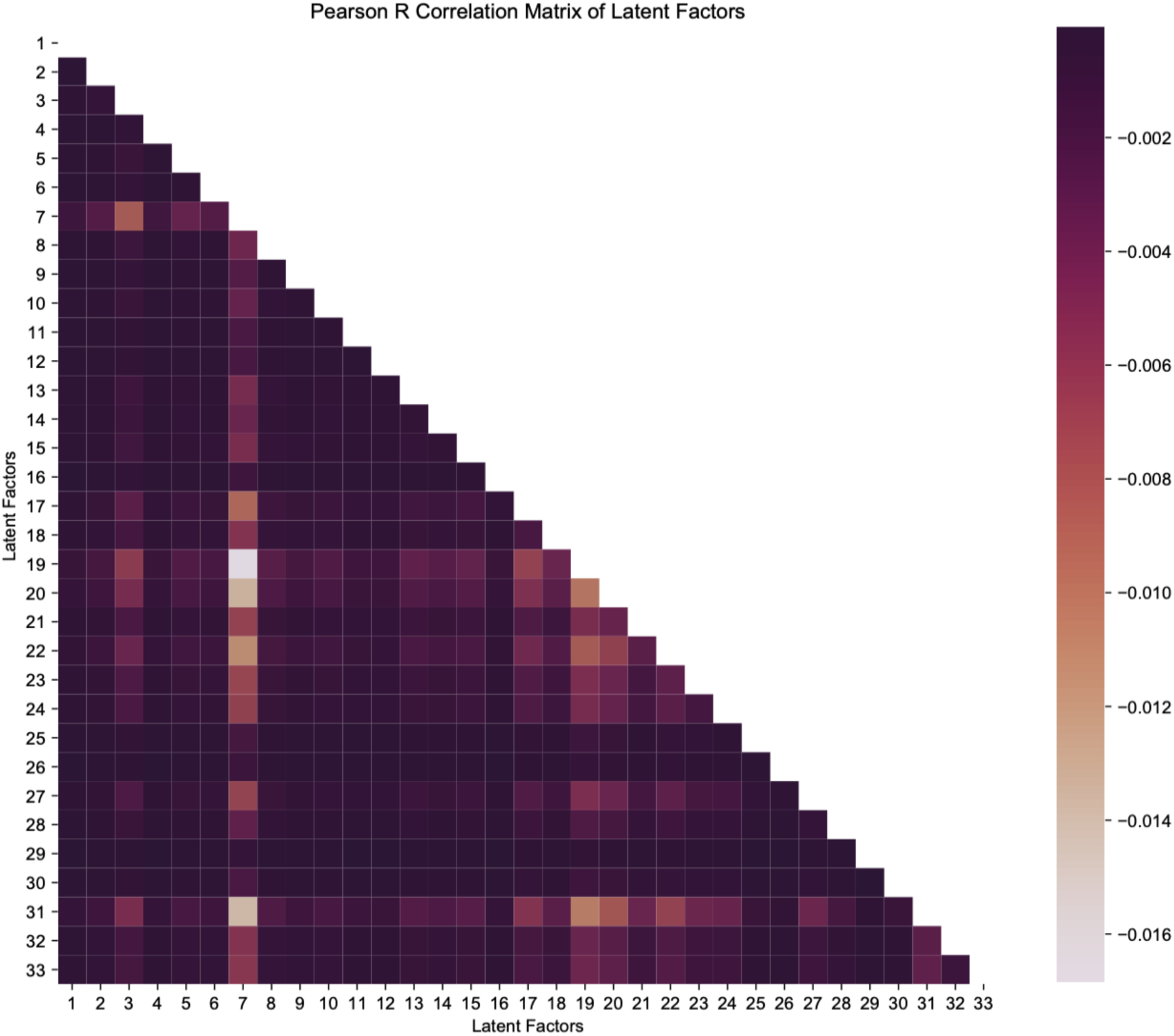
Pearson correlation of latent factors. We observed low correlations between latent factors across phenotype, individual, and time at Vanderbilt.

## SUPPLEMENTAL TABLES

**Supplemental Table S1:** Phecodes associated with preterm birth in an African American cohort passing multiple testing correction at Vanderbilt University.

| **Phecode** | **Description** | **OR** | **P-value** | **-log10p** | **Category** |
| --- | --- | --- | --- | --- | --- |
| 653 | Problems associated with amniotic cavity and membranes | 5.46 | 3.07E-76 | 75.51 | pregnancy complications |
| 642.1 | Preeclampsia and eclampsia | 5.33 | 8.23E-55 | 54.08 | pregnancy complications |
| 636.1 | Threatened premature labor | 4.95 | 2.03E-36 | 35.69 | pregnancy complications |
| 642 | Hypertension complicating pregnancy, childbirth, and the puerperium | 2.53 | 1.95E-26 | 25.71 | pregnancy complications |
| 636.2 | Early onset of delivery | 82.83 | 2.07E-26 | 25.68 | pregnancy complications |
| 636.8 | Cervical incompetence | 13.41 | 5.22E-26 | 25.28 | pregnancy complications |
| 401.1 | Essential hypertension | 3.39 | 2.34E-24 | 23.63 | circulatory system |
| 635.2 | Antepartum hemorrhage, abruptio placentae, and placenta previa | 5.94 | 3.99E-22 | 21.40 | pregnancy complications |
| 250.2 | Type 2 diabetes | 3.94 | 8.96E-18 | 17.05 | endocrine/metabolic |
| 655 | Known or suspected fetal abnormality affecting management of mother | 1.65 | 1.17E-15 | 14.93 | pregnancy complications |
| 634 | Miscarriage; stillbirth | 3.58 | 2.31E-15 | 14.64 | pregnancy complications |
| 649.1 | Diabetes or abnormal glucose tolerance complicating pregnancy | 2.02 | 2.64E-12 | 11.58 | pregnancy complications |
| 250.1 | Type 1 diabetes | 5.95 | 6.11E-11 | 10.21 | endocrine/metabolic |
| 250.3 | Insulin pump user | 8.93 | 5.49E-09 | 8.26 | endocrine/metabolic |
| 278.11 | Morbid obesity | 2.05 | 1.29E-07 | 6.89 | endocrine/metabolic |
| 649 | Other conditions or status of the mother complicating pregnancy, childbirth, or the puerperium | 1.41 | 3.93E-07 | 6.41 | pregnancy complications |
| 276.5 | Hypovolemia | 2.73 | 6.32E-07 | 6.20 | endocrine/metabolic |
| 428.1 | Congestive heart failure (CHF) NOS | 5.55 | 1.90E-06 | 5.72 | circulatory system |
| 401.22 | Hypertensive chronic kidney disease | 12.51 | 3.56E-06 | 5.45 | circulatory system |
| 512.7 | Shortness of breath | 2.39 | 4.19E-06 | 5.38 | respiratory |
| 585.3 | Chronic renal failure [CKD] | 12.17 | 4.60E-06 | 5.34 | genitourinary |
| 625 | Pain and other symptoms associated with female genital organs | 2.32 | 4.62E-06 | 5.34 | genitourinary |
| 783 | Fever of unknown origin | 2.21 | 4.94E-06 | 5.31 | symptoms |
| 269 | Proteinuria | 6.45 | 6.16E-06 | 5.21 | endocrine/metabolic |
| 585.32 | End stage renal disease | 11.12 | 1.17E-05 | 4.93 | genitourinary |
| 585.1 | Acute renal failure | 3.52 | 1.37E-05 | 4.86 | genitourinary |
| 285 | Other anemias | 2.27 | 2.40E-05 | 4.62 | hematopoietic |
| 272.1 | Hyperlipidemia | 4.02 | 3.19E-05 | 4.50 | endocrine/metabolic |

**Supplemental Table S2:**
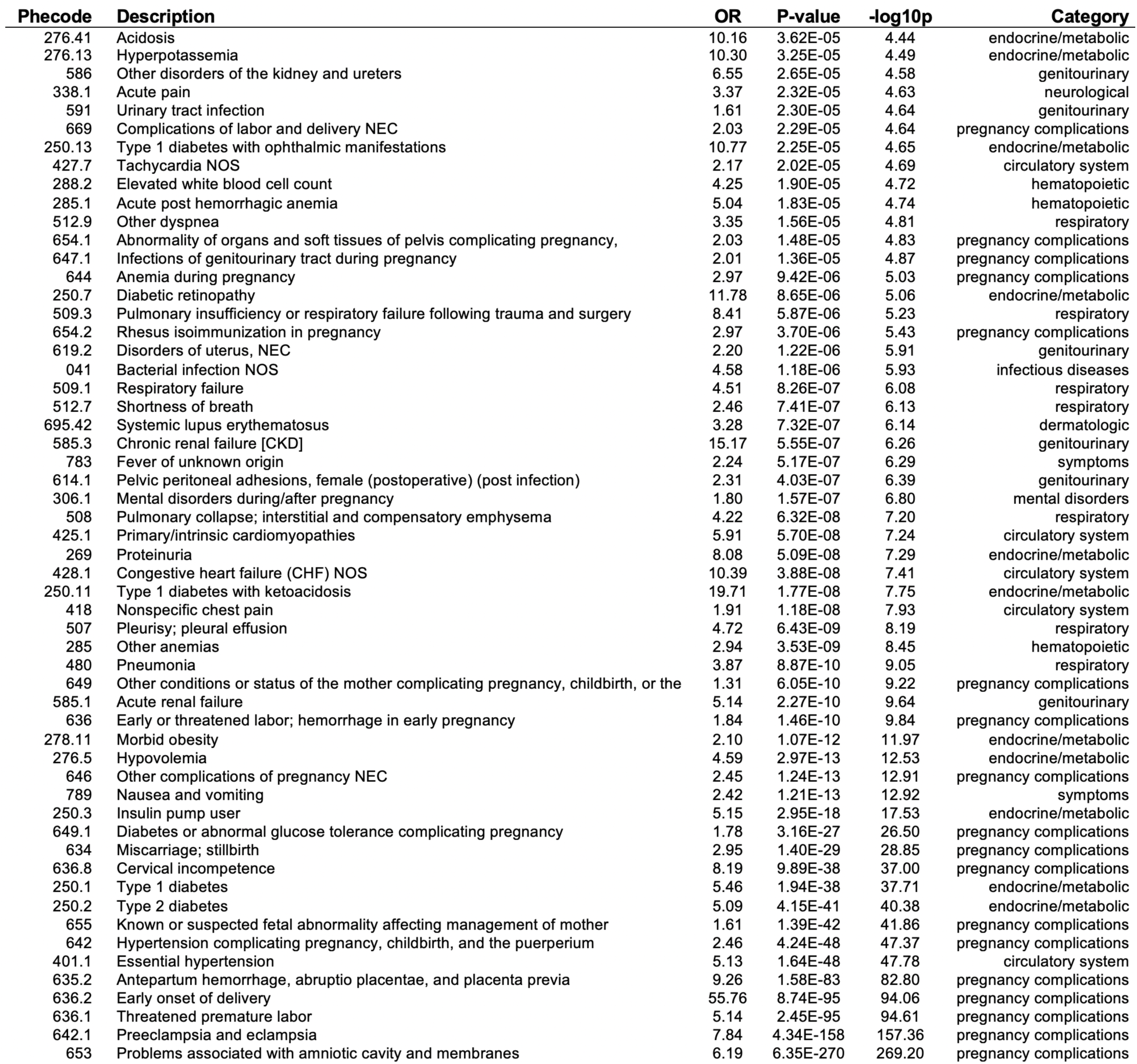
Phecodes associated with preterm birth in a Caucasian cohort passing multiple testing correction at Vanderbilt University.

**Supplemental Table S3:**
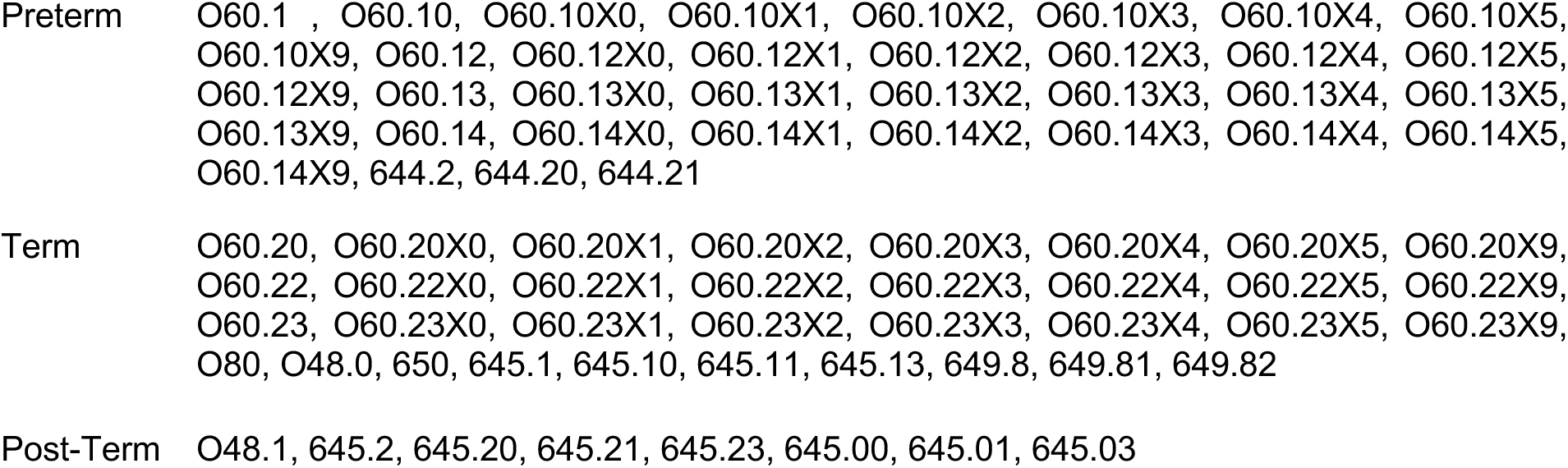
ICD-9 delivery codes excluded prior to creating longitudinal disease tensor.

**Supplemental Table S4:** Top phecodes for latent factor 6.

| **phecode** | **weight** | **description** | **phenotype group** |
| --- | --- | --- | --- |
| 649.1 | 34.5 | Diabetes or abnormal glucose tolerance complicating pregnancy | pregnancy complications |
| 250 | 16.5 | Diabetes mellitus | endocrine/metabolic |
| 250.2 | 12.1 | Type 2 diabetes | endocrine/metabolic |
| 250.1 | 8.8 | Type 1 diabetes | endocrine/metabolic |
| 250.3 | 6.0 | Insulin pump user | endocrine/metabolic |
| 250.7 | 1.0 | Diabetic retinopathy | endocrine/metabolic |
| 250.11 | 0.9 | Type 1 diabetes with ketoacidosis | endocrine/metabolic |
| 251.1 | 0.8 | Hypoglycemia | endocrine/metabolic |
| 250.13 | 0.7 | Type 1 diabetes with ophthalmic manifestations | endocrine/metabolic |
| 250.12 | 0.6 | Type 1 diabetes with renal manifestations | endocrine/metabolic |

**Supplemental Table S5:** Top phecodes for latent factor 13.

| **phecode** | **weight** | **description** | **group** |
| --- | --- | --- | --- |
| 244 | 30.1 | Hypothyroidism | endocrine/metabolic |
| 244.4 | 27.8 | Hypothyroidism NOS | endocrine/metabolic |
| 244.1 | 4.0 | Secondary hypothyroidism | endocrine/metabolic |
| 245 | 3.9 | Thyroiditis | endocrine/metabolic |
| 245.2 | 2.5 | Chronic thyroiditis | endocrine/metabolic |
| 245.21 | 2.5 | Chronic lymphocytic thyroiditis | endocrine/metabolic |
| 244.2 | 1.8 | Acquired hypothyroidism | endocrine/metabolic |
| 193 | 1.2 | Thyroid cancer | neoplasms |
| 253 | 0.6 | Disorders of the pituitary gland and its hypothalamic control | endocrine/metabolic |
| 253.1 | 0.4 | Pituitary hyperfunction | endocrine/metabolic |

